# The Generalized 3+3 (G3+3) Design for Phase I Dose-Finding Trials

**DOI:** 10.1101/2024.08.18.24312178

**Authors:** Yuan Ji, Yunxuan Zhang, Andrew Ji

**Affiliations:** Department of Public Health Sciences, The University of Chicago, IL; The University of Chicago Laboratory Schools, IL

## Abstract

**PURPOSE:** We propose and demonstrate the feasibility and desirability of a novel model-free dose-finding design for phase I clinical trials.

**METHODS:** The Generalized 3+3 (G3+3) design uses a set of simple rules summarized as follows: For 3 or 6 patients at a dose, apply the 3+3 design for making dosing decisions. For other numbers, if the observed toxicity rate (OTR) is less than 0.2, escalate to the next higher dose; if the OTR is greater than 0.29, de-escalate to the next lower dose; otherwise, stay at the current dose.

**RESULTS:** The G3+3 design is the only design that can replicate the decisions of the 3+3 design for 3 or 6 patients among the popular designs compared like BOIN and i3+3. G3+3 generates desirable decisions when the number of patients treated is not 3 or 6, like the popular designs. Computer simulation verifies the superior operating characteristics of the G3+3 design.

**CONCLUSION:** The G3+3 design generalizes the popular 3+3 design so that desirable decisions can be made for any number of patients at a dose. G3+3 does not rely on statistical models, is simple and transparent, and can be implemented without a software tool. Therefore, it is expected to facilitate and enhance modern phase I dose-finding trials and early-phase drug development.

## Introduction

Classical phase I trials aim to identify the maximum tolerated dose (MTD). A statistical design provides dose assignment decisions continuously to the next patients enrolled in the trial based on the observed toxicity outcomes from previously enrolled patients, usually recorded as a binary dose-limiting toxicity endpoint (DLT). The last three decades have seen incredible advancements in statistical designs for phase I dose-finding trials. A major movement (Ji & Yuan, 2021; Kurzrock et al., 2021) is to utilize statistical models rather than model-free rules like those in the 3+3 design (Storer, 1989). Pioneered by the continual reassessment method (CRM, O’Quigley et al., 1990), a large number of novel designs (Guo et al., 2017; Ji et al., 2010; Ji & Wang, 2013; Liu et al., 2020; Liu & Yuan, 2015; O’Quigley et al., 1990) using statistical models have been proposed, all of which rely on probability models to estimate dose toxicity probabilities and make dosing decisions. However, gradually the newer designs resort to simpler models aiming to make the designs user-friendly. For example, a class of model-assisted designs like mTPI-2 (Guo et al., 2017) and BOIN (Liu & Yuan, 2015) highlights the philosophy of using simplified statistical models, the latter receiving FDA’s fit-for-purpose designation in 2021. More recently, a model-free design called i3+3 (Liu et al., 2020) seems to suggest that a smart dose-finding algorithm that does not use statistical models might achieve similar superior performance to model-dependent designs. Duan et al., (2024) summarize the development of new dose-finding designs as a spiral evolution that progresses from model-free designs like 3+3, to model-based designs like CRM, to model-assisted designs like BOIN, and then back to model-free designs like i3+3.

Such a spiral evolution is confusing. For example, one may ask, does an MTD-seeking phase I dose-finding design need statistical models? Why is 3+3 not good? We address both questions in this work. Regarding the second question, the main issue with the 3+3 design is that it cannot deal with cases when the number of patients treated at a dose is not 3 or 6. However, the decisions for 3 or 6 patients based on the 3+3 design are considered the gold standard in the clinical community and have been upheld in numerous real-world trials. For the first question, we argue that if the objective to identify the MTD and testing the safety profiles of a few ascending dose, an effective dose-finding design does not need to use a probability model. If other objectives such as estimating dose-response relationship are of interests, model-based designs are needed. We show that by generalizing the idea of 3+3 beyond 3 or 6 patients, model-free and rule-based designs may perform equally well as recently developed novel designs and better than the 3+3 design.

To this end, we propose a new design called the Generalized 3+3 (G3+3) design. The G3+3 design does not rely on a probability model for decision making and is based on a set of simple and intuitive decision rules. The implementation of G3+3 can be easily carried out in practice by both statisticians and clinicians. Unlike 3+3, the G3+3 design provides desirable decisions for any number of patients and these decisions can be pre-tabulated for assessment. We first explain the basic idea of G3+3 next.

## The G3+3 Design

### Algorithm

Suppose at any moment a trial is testing a dose by assigning *n* patients to the dose. Let us call the dose “the current dose.” Suppose among the *n* treated patients *y* of them experience DLT. Then the observed toxicity rate (OTR) is simply 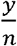. For example, given *n* = 3, *y* = 1, the OTR is 1/3.

Investigators specify two values *L* and *H*. A dose with toxicity probability lower than *L* is considered below the MTD, higher than *H* above the MTD, and between *L* and *H* close to the MTD. We propose the following dose escalation rules following Ivanova and Flournoy (Ivanova et al., 2007).

1. If the OTR is below *L*, escalate (E) and treat patients at the next higher dose;
2. If the OTR is above *H*, de-escalate (D) and treat patients at the next lower dose;
3. If the OTR is between *L* and *H* (including *L* and *H*), stay (S) and treat patients at the same current dose.

The decisions E, D, and S are called up-and-down decisions that allocate future patients to the adjacent dose above the current dose, below the current dose, and at the current dose, respectively. The default G3+3 design consists of two sets of *L* and *H* values.

⍰ When *n* is equal to or less than 3, *L*= 0.2 and *H* = 1/3;
⍰ When *n* is greater than 3, *L*= 0.2 and *H* = 0.29.

The G3+3 design continues to escalate (E), de-escalate (D), or stay (S) based on rules 1-3 until the trial ends.

### Decisions

#### Number of patients is 3 or 6

We illustrate the decisions based on the proposed G3+3 design. First, we examine its decisions when 3 or 6 patients are treated at a dose in Table 1 and compare them with four other designs, 3+3, i3+3, BOIN, and mTPI-2. G3+3 is the only design in the list that can replicate the 3+3 design’s decisions.

**Table 1.**
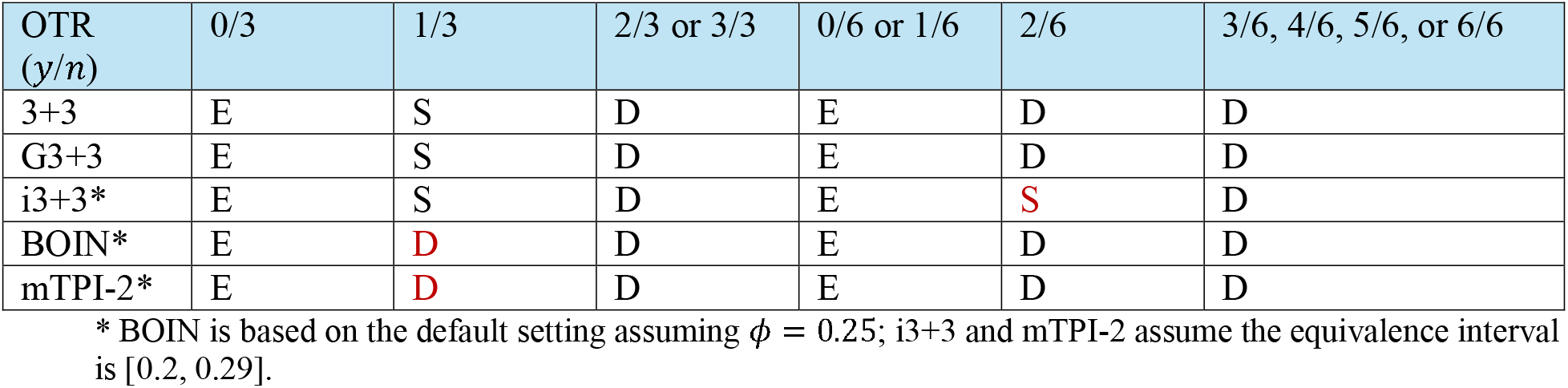
Dosing decisions of the 3+3, G3+3, i3+3, BOIN, and mTPI-2 designs for 3 or 6 patients. The decisions different from the 3+3 design are highlighted in red. Clearly, only G3+3 matches 3+3 in all cases. Here E, S, and D mean to escalate to the next higher dose, stay at the current dose, and de-escalate to the next lower dose, respectively.

| OTR<br>($y/n$) | 0/3 | 1/3 | 2/3 or 3/3 | 0/6 or 1/6 | 2/6 | 3/6, 4/6, 5/6, or 6/6 |
| --- | --- | --- | --- | --- | --- | --- |
| 3+3 | E | S | D | E | D | D |
| G3+3 | E | S | D | E | D | D |
| i3+3* | E | S | D | E | **S** | D |
| BOIN* | E | **D** | D | E | D | D |
| mTPI-2* | E | **D** | D | E | D | D |

\* BOIN is based on the default setting assuming $\phi = 0.25$ ; i3+3 and mTPI-2 assume the equivalence interval is $[0.2, 0.29]$ .

**A unique feature of the 3+3 design** is that it stays (S) at the current dose when one out of three patients experiences DLT but de-escalates (D) to the next lower dose when two out of six patients experience DLT. In both cases, the OTR is 1/3. The decision D at “two out of six” implies that 3+3 considers the toxicity rate 1/3 to be excessive, and therefore the design targets a toxicity rate lower than 1/3. However, when the data is “one out of three” the 3+3 design recognizes the small sample size of 3 and is willing to give the dose another try by staying (S) at the dose.

These two different 3+3 decisions highlight the underlying statistical thinking behind the 3+3 design and partly justify its popularity among the clinical society. Recently developed designs like mTPI-2 (Guo et al., 2017) and BOIN (Liu & Yuan, 2015) are not able to distinguish the differences in these two cases and therefore would make the same decision for both cases, de-escalate (D). The i3+3 design (Liu et al., 2020) tries to differentiate the two cases with a smart rule but generates an undesirable decision (S) when the OTR is 2/6 (Table 1). In summary, only G3+3 can replicate the decisions of the 3+3 design for three or six patients.

#### Number of patients is not 3 or 6

The G3+3 design can generate a decision for any number of patients based on the proposed algorithm. For example, Figure 1 presents a decision table for the 3+3, G3+3, i3+3, and BOIN designs for up to 9 patients at a dose. When the number of patients is not 3 or 6, the decisions of the G3+3 design are identical to those of the i3+3 and BOIN designs. In Figure 1 the decision DU means to de-escalate (D) and that the current dose is unacceptable (U) due to the overly toxic OTR. A dose with DU decision will be removed from the trial, as well as other doses that are higher than the dose. We will discuss how DU is generated next.

**Figure 1.**
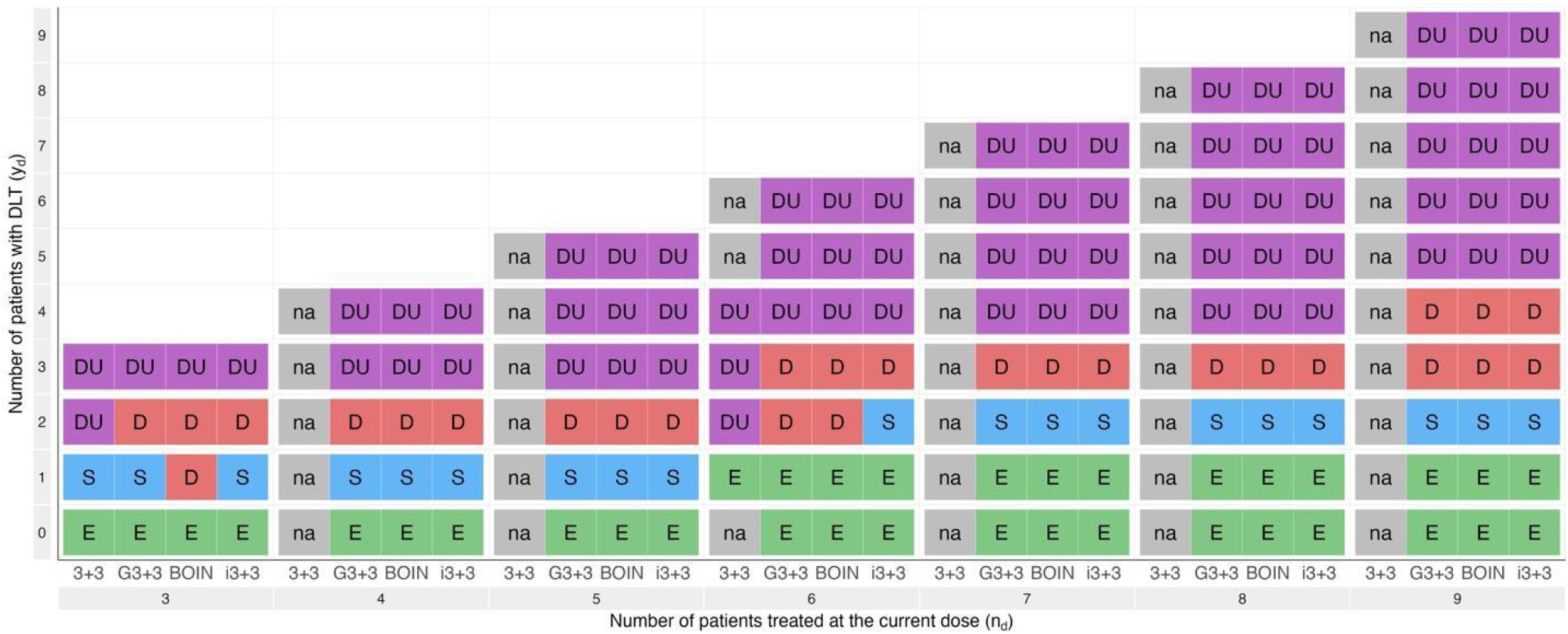
Dosing decisions for the 3+3, G3+3, BOIN, and i3+3 designs for up to 9 patients. Here E, S, and D mean to escalate to the next higher dose, stay at the current dose, and de-escalate to the next lower dose, respectively. Decision DU means D and that the current dose is excluded from the trial due to unacceptable (U) toxicity. The 3+3 design only provides decisions for 3 or 6 patients and therefore shows “na” at places when the number of patients is not 3 or 6. G3+3 is the only design that replicates all the 3+3 decisions at 3 and 6 patients and the BOIN and i3+3 decisions at other numbers. The BOIN and i3+3 designs are based on the default setting where the target toxicity probability is 0.25, and the equivalence interval for i3+3 is [0.2, 0.3].

### Safety Rule, Special Cases, and MTD Selection

#### Safety Rule

In Figure 1, some decisions are marked as DU. For example, when three out of three patients have DLT at a dose, the decision in Figure 1 is DU for all the designs. The letter U is generated based on a safety rule proposed in the mTPI design (Ji et al., 2010) and later adopted widely by other designs like mTPI-2, BOIN, and i3+3. To facilitate practical application of the G3+3 design, Table 2 lists all the DU decisions for different numbers of patients treated at a dose.

**Table 2.**
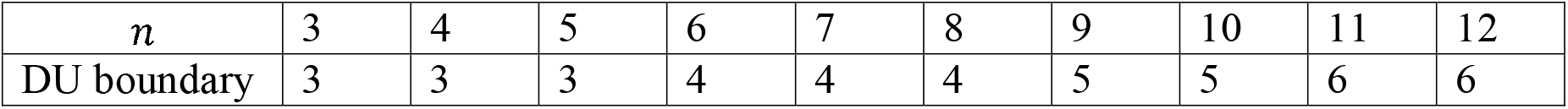
For *n* patients treated at a dose, if the number of patients *y* experiencing DLT is equal to or greater than the DU boundary, the dose is marked as DU, i.e., one must de-escalate to the next lower dose and remove the dose (and any dose higher than the dose) from the trial. Statistically, the boundary is calculated as follows: given *y* and *n* at a dose, if the posterior probability that the toxicity rate of the dose is larger than 0.25 is greater than 0.95, the dose is marked as DU. The posterior probability is based on the binomial likelihood and a beta(1,1) distribution.

#### Special Cases

When the lowest dose has a D decision (not DU) or when the highest dose has an E decision, change either decision to S. These two special cases are needed since the original decisions cannot be executed. Decisions when *n* = *1* or *2* are not listed since they are usually special cases up to the investigator’s discretion. For example, when *n* = *1*, and the patient experiences DLT, sometimes, the decision is to stay (S) and enroll two more patients.

#### MTD Selection

The G3+3 design uses a simple and effective rule to select the MTD at the end of the trial when dose finding is complete and the task is to select a dose as the MTD.

⍰ After removing doses based on the safety rule, at the end of the trial, G3+3 selects the highest tested dose for which the decision is not D.
⍰ If no tested doses have a decision D (meaning their decisions are either E or S), select the highest tested dose as the MTD.
⍰ If the lowest dose has a decision D, no dose is selected as the MTD.

The MTD selection rule is another simplification compared with the popular designs like i3+3, BOIN and mTPI-2, which apply an isotonic regression at the end based on a beta-binomial model. Isotonic regression is a complex mathematical operation that requires computer programs and software. In contrast, the proposed simple MTD selection procedure for G3+3 can be carried out by hand.

### Cohort Size and Sample Size

The G3+3 design can generate a decision for any number of patients at a dose. Hence a trial under G3+3 may enroll any number of patients in a cohort. Conventionally, the cohort size is 3 although it needs not be. One could also vary the cohort size during a trial. By default, we recommend using a cohort size of 3.

If the lowest dose is marked as DU, the trial stops and no dose is selected as the MTD. Otherwise, continuously apply the G3+3 algorithm until a pre-specified max number is reached, such as 6 times the number of doses. For example, if there are five doses in the trial, the sample size could be 30. Additionally, one may apply a rule that stops a trial if a prespecified number of patients, say 9 or 12, have been treated at any dose.

## Implementation Algorithm of G3+3

To implement the G3+3 design, we propose a simple procedure of three steps.

### Step 1 [Dose Escalation]

For 3 or 6 patients at the current dose, apply the decisions of the 3+3 design. For other numbers, if the OTR is less than 0.2, E; if the OTR is greater than 0.29, D; otherwise, S. This procedure is equivalent to the proposed G3+3 algorithm but simpler to remember and implement in practice.

### Step 2 [Safety Rule]

Apply the DU boundary (Table 2) to remove overly toxic doses throughout the trial.

### Step 3 [MTD Selection]

A trial is stopped either when a prespecified sample size is reached or when no dose is left. In the former case, apply the MTD selection rule to find an MTD. In the latter case, no dose is selected.

To facilitate the implementation of the G3+3 design, we attach a supplemental document (Appendix) that can be inserted into a phase I trial protocol using the G3+3 design. Implementation of the G3+3 design requires no computer program. Furthermore, the simulation results in Appendix can be repeatedly utilized for conventional dose-finding trials to facilitate practical application.

## Simulation

We compare G3+3 to CRM, i3+3 and BOIN as the latter three designs have all shown to exhibit superior operating characteristics to the 3+3 design (Iasonos & O’Quigley, 2014; Liu et al., 2020; Liu & Yuan, 2015). The design parameters for the four designs are summarized in the Appendix (Table A.1). The G3+3 design is expected to generate similar patient allocation decisions to these designs because its decisions are similar as shown in Figure 1. We summarize patient allocation in the Appendix to be concise. Here, we focus on the performance of G3+3 in MTD selection since it uses a new and simpler MTD selection rule, as opposed to the popular model-based MTD selection based on Bayesian models and isotonic transformation.

Figure 2 presents the design performance in terms of the percentage of trials that select the correct MTD (PCS), a dose over the MTD (POS), or a dose under the MTD (PUS). The simulation scenarios (Table 3) are adopted from (Zhou et al., 2021) and also used in the FDA review of the BOIN design for its fit-for-purpose designation.

**Table 3.**
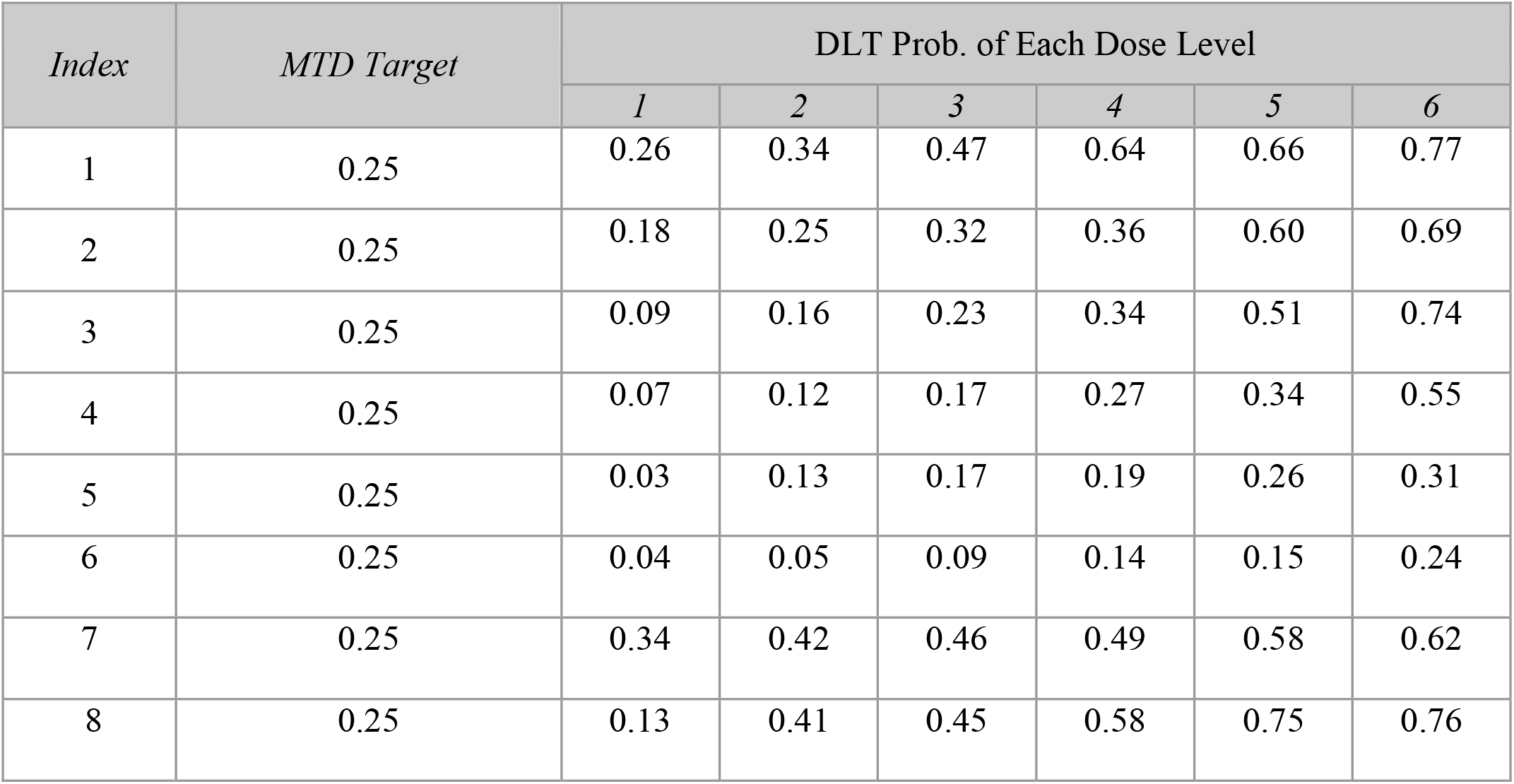
A list of eight scenarios with true toxicity probabilities for six doses per scenario.

**Figure 2.**
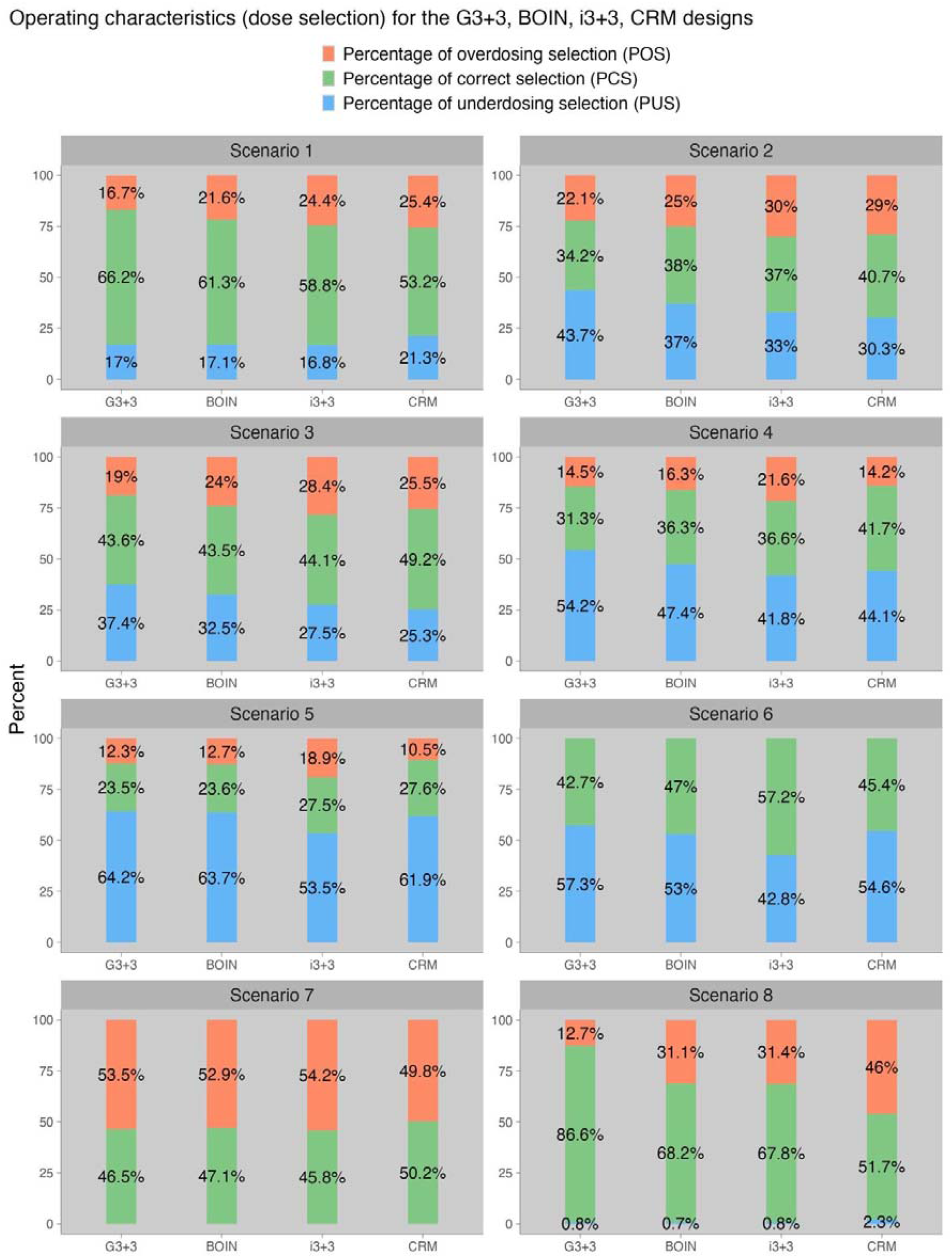
Operating characteristics in terms of MTD selection for the G3+3, BOIN, i3+3 and CRM designs.

Regarding PCS, the CRM design is overall the best with the highest PCS in most scenarios, while G3+3’s performance is comparable to i3+3 and BOIN. G3+3 outperforms CRM, i3+3 and BOIN in Scenario 8, where the true MTD is Dose 1. This scenario might resemble practical trials for novel cancer therapies that rarely induce DLTs at any dose level. In addition, compared to i3+3 and BOIN, G3+3 has relatively low POS. This observation indicates that the proposed simple MTD selection rules are more conservative and desirable in controlling the risk of overdosing than those designs like i3+3 and BOIN based on isotonic regression. To be more specific, the proposed simple MTD selection rule will always select the highest dose for which the decision is either S or E, while the rule based on the isotonic regression will occasionally select a dose for which the decision is D. Therefore, the new MTD selection rule might be preferred due to its safety and simplicity. The model-based design CRM shows desirable performance. However, it requires infrastructure and expertise to implement in practice -- a tradeoff to be balanced in practice.

## Discussion

Phase I trial designs have gone through multiple iterations of refinement. For more than three decades, focus has been put on the development of probability model-based designs to improve classic rule-based designs like 3+3. However, over time the models became simpler and the recent development of the i3+3 design seems to suggest model-free designs might be just as efficient as model-dependent designs.

Numerous publications have demonstrated the inferiority of the 3+3 design, mostly focusing on its poor performance in simulation. However, we argue the main issue of the 3+3 design is its inability to make dosing decisions when the number of patients treated at a dose is not 3 or 6, resulting in premature trial stopping and lack of flexibility in practice. For example, 3+3 cannot handle cases of over-enrollment or dropouts that may result in the number of patients at a dose to be neither 3 or 6. In these cases, ad-hoc decisions are needed which result in protocol deviation. Nevertheless, the 3+3 decisions for 3 or 6 patients have been widely accepted as a standard and verified in numerous trials. The question is then how to preserve these decisions and at the same time extend them to cases when the number of patients is not 3 or 6.

To this end, we have shown that the G3+3 design can answer the question. By simply comparing the observed toxicity rate at a dose to a pair of decision boundaries denoted as *L* and *H*, and judiciously using a different a pair of (*L, H*) when the number of patients is less than or equal to 3 versus greater than 3, one can generate the exact set of decisions in 3+3 as well as those of the popular designs like i3+3 and BOIN when the number of patients is not 3 or 6. In practice, in order to apply the default G3+3 design, a clinician simply needs to follow the three steps in the Implementation Algorithm. These steps are straightforward and do not require any advanced statistics or computing. The statistical section for trial protocol and simulation results provided in Appendix are expected to greatly facilitate adoption of G3+3 in practice. For the vast majority of phase I trials, the default G3+3 design of setting *L* = 0.2 and *H* = 0.29 for *n* > 3 and *L*= 0.2 and *H* = 1/3 for *n* ≤ *3* is sufficient as the toxicity probability of the MTD is usually within the range of 0.2 and 0.3.

Lastly, the development of G3+3 does not imply that any dose-finding designs can be free of probability models. We only consider the type of trial that uses DLT as the primary endpoint. In modern drug development, dose-finding trials have become more complex considering that attempts are made to capture various endpoints like efficacy and patient reported outcomes. Statistical models can always help explain variabilities in the data, mitigate biased decisions, and improve trial efficiency. In the simplest case of DLT-based dose finding, especially with small sample size per dose, model-free designs like G3+3 are a special case.

## Data Availability

There is no data in the article since it is about a statistical design for phase I clinical trials.

