## Appendix for "The Generalized 3+3 (G3+3) Design for Phase I Dose-Finding Trials"

### **Appendix: Protocol Writing Using the G3+3 Design**

### **X. Statistical Method**

#### **X.X Statistical Design – The G3+3 design**

There are six dose levels in the trial. The maximum sample size is 36^[1]^.

The G3+3 design consists of three parts, dose escalation, safety rule, and MTD selection.

**Dose Escalation** Suppose at any moment a trial is testing a dose with a total of $n$ patients assigned to the dose. Let us call the dose “the current dose.” Suppose among $n$ patients $y$ of them experience dose-limiting toxicity (DLT). Define the observed toxicity rate (OTR) as the ratio $\frac{y}{n}$. The G3+3 design compares the OTR $\frac{y}{n}$ with a low value $L$ and a high value $H$. A dose with toxicity probability lower than $L$ is considered below the MTD, higher than $H$ above the MTD, and between $L$ and $H$ close to the MTD. The G3+3 design uses the following dose escalation rules.

1. If the OTR is below $L$, escalate (E) and treat patients at the next higher dose;
2. If the OTR is above $H$, de-escalates (D) and treat patients at the next lower dose;
3. If the OTR is between $L$ and $H$ (including $L$ and $H$), stay (S) and treat patients at the same current dose.

The G3+3 design consists of two sets of $L$ and $H$ values. When $n$ is equal to or less than 3, $L=0.2$ and $H=1/3$; when $n$ is greater than 3, $L=0.2$ and $H=0.29$. The trial continues to escalate (E), de-escalate (D), or stay (S) based on the rules 1-3 until the end.

**Safety Rule** The G3+3 design applies a safety rule to remove doses that are deemed overly toxic from the trial. When $n$ number of patients are tested at the current dose and the observed DLT number $y$ reaches or exceeds the DU boundary in Table A.1, the current dose and all the higher doses are removed from the trial permanently.

Table A.1 For $n$ patients treated at a dose, if the number of patients $y$ experiencing DLT is equal to or greater than the DU boundary, the dose is marked as DU, i.e., one must de-escalate to the next lower dose and remove the dose (and any dose higher than the dose) from the trial. Statistically, the boundary is calculated as follows: given $y$ and $n$ at a dose, if the posterior probability that the toxicity rate of the dose is larger than 0.25 is greater than 0.95, the dose is marked as “DU”. The posterior probability is based on the binomial likelihood and a beta(1,1) distribution.

| $n$ | 3 | 4 | 5 | 6 | 7 | 8 | 9 | 10 | 11 | 12 |
| --- | --- | --- | --- | --- | --- | --- | --- | --- | --- | --- |
| DU boundary | 3 | 3 | 3 | 4 | 4 | 4 | 5 | 5 | 6 | 6 |

The G3+3 decisions are listed in Figure A.1 below for up to $n=12$ patients. However, the G3+3 design can generate a decision for any number of $n$ based on the above rules.

Figure A. 1 The dosing decisions of the G3+3 design using the default setting. Here E, S, and D mean to escalate to the next higher dose, stay at the current dose, and de-escalate to the next lower dose, respectively. Decision DU means D and that the current dose is excluded from the trial due to Unacceptable toxicity.

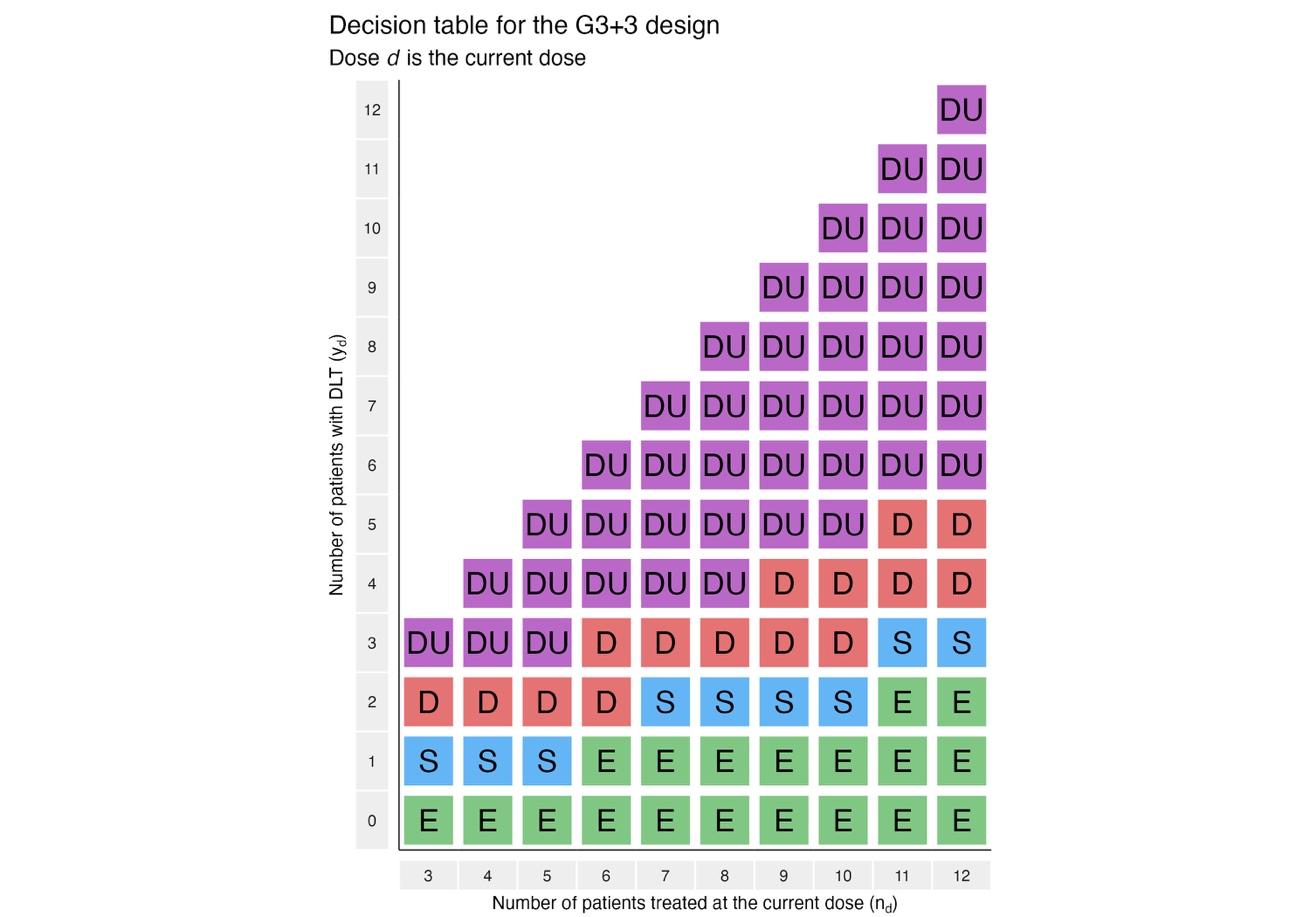

When the lowest dose has a “D” decision (not “DU”), change to “S”. When the highest dose has an “E” decision, change to “S”. These two special cases are needed since the original decisions cannot be executed.

**MTD Selection** The G3+3 design uses a simple and effective rule to select the MTD at the end of the trial. After removing doses based on the safety rule, at the end of the trial, G3+3 selects the highest tested dose for which the decision is not D. If no tested doses have a decision D (meaning their decisions are either E or S), select the highest tested dose as the MTD. If the lowest dose has a decision D, no dose is selected as the MTD.

To implement the G3+3 design, follow the next three steps.

**Step 1 [Dose Escalation]** For 3 or 6 patients at the current dose, apply the decisions of the 3+3 design. For other numbers, if the OTR is less than 0.2, E; if the OTR is greater than 0.29, D; otherwise, S. This procedure is equivalent to the proposed G3+3 algorithm but simpler to remember and implement.

**Step 2 [Safety Rule]** Apply the DU boundary to remove overly toxic doses throughout the trial.

**Step 3 [MTD Selection]** A trial is stopped either when the maximum sample size is reached or no dose is left. In the former case, apply the MTD selection rule to find an MTD. In the latter case, no dose is selected.

**X.Y Simulation Results of G3+3**

We conduct simulations to compare G3+3 design with CRM, i3+3 and BOIN designs (Liu et al., 2020; Liu & Yuan, 2015; O'Quigley et al., 1990). The simulation settings and results are presented next.

#### Scenario list

A total of six simulated scenarios are listed in the following Table A.2, with 5,000 simulations for each scenario. These scenarios are adopted from Zhou et al. (Zhou et al., 2021).

Table A.2． A list of eight scenarios with true toxicity probabilities for six doses per scenario.

| *Index* | *MTD Target* | DLT Prob. of Each Dose Level | | | | | |
| --- | --- | --- | --- | --- | --- | --- | --- |
|  |  | *1* | *2* | *3* | *4* | *5* | *6* |
| 1 | 0.25 | 0.26 | 0.34 | 0.47 | 0.64 | 0.66 | 0.77 |
| 2 | 0.25 | 0.18 | 0.25 | 0.32 | 0.36 | 0.60 | 0.69 |
| 3 | 0.25 | 0.09 | 0.16 | 0.23 | 0.34 | 0.51 | 0.74 |
| 4 | 0.25 | 0.07 | 0.12 | 0.17 | 0.27 | 0.34 | 0.55 |
| 5 | 0.25 | 0.03 | 0.13 | 0.17 | 0.19 | 0.26 | 0.31 |
| 6 | 0.25 | 0.04 | 0.05 | 0.09 | 0.14 | 0.15 | 0.24 |
| 7 | 0.25 | 0.34 | 0.42 | 0.46 | 0.49 | 0.58 | 0.62 |
| 8 | 0.25 | 0.13 | 0.41 | 0.45 | 0.58 | 0.75 | 0.76 |

#### Design list

Table A.3 lists the four designs and their configurations.

Table A.3． Four designs and their configurations in the simulations.

|  | G3+3 | i3+3 | BOIN | CRM |
| --- | --- | --- | --- | --- |
| Target DLT rate | 0.25 | 0.25 | 0.25 | 0.25 |
| ($L, H$) values | $[0.2, \frac{1}{3}]\text{ for }n\leq3$  $[0.2,0.29]\text{for }n>3$ | $[0.2,0.3]$ | $\left[ 0.198,0.297 \right)$ | N/A |
| Sample size | 36 | 36 | 36 | 36 |
| Cohort size | 3 | 3 | 3 | 3 |
| Model | N/A | N/A | $y_{d}\sim Bin(n_{d},p_{d})$ | $p_{d}=p_{0d}^{\exp\left( \alpha\right)}$ |
| Prior | N/A | N/A | $p_{d}\sim\frac{1}{3}I\left( p_{d}=0.25 \right)+\frac{1}{3}I\left( p_{d}=0.15 \right)+\frac{1}{3}I(p_{d}=0.35)$ | $\alpha\sim LogNormal$*(0,2)*  $Skeleton \left( p_{01},\ldots,p_{06} \right)$^*^ |
| Dose-skipping | No | No | No | No |
| Starting dose | 1 | 1 | 1 | 1 |
| Overdose control rule | $Pr(p_{d}>0.25\left\vert data \right)>0.95$ | | | N/A |
| Stopping rule | $Pr(p_{1}>0.25\left\vert data \right)>0.95$ | | | |

Notation: $p_{d}$ denotes the true DLT probability of dose level *d*; $y_{d}$ denotes the number patients experienced DLTs at dose level *j*; $n_{d}$ denotes the number of patients treated at dose level *j*. * In our simulation, we used skeletons ${(p}_{01},\ldots,p_{06})=$(0.062, 0.140, 0.25, 0.376, 0.502, 0.615) for a target DLT probability 0.25, obtained using the method of Lee and Cheung (2009) from R package “dfcrm”.

#### Simulation results

The operating characteristics are demonstrated in stacked bar plots arranged by scenarios. Three stacked bar plots present the four designs in the design list, respectively. In each scenario-specific panel, the trial-specific summary statistics are reported, mainly from two aspects: MTD Selection (Figure A.2, which is the same as Figure 2 in the main paper), and Patient Allocation (Figure A.3). Specifically, they are

- MTD Selection
  - Percentage of Correct Selection (PCS): The percentage of all simulated trials selecting the true MTD, defined as the percentage of simulated trials that correctly select the true MTD across all the trials. The higher the value, the better the design.
  - Percentage of Overdosing Selection (POS): The percentage of all simulated trials selecting the dose levels above the true MTD, which is defined by the percentage of simulated trials that select a dose higher than the true MTD at the end of the trial across all the simulated trials. The lower the value, the better the safety of the design.
  - Percentage of Underdosing Selection (PUS): The percentage of all simulated trials selecting the dose levels below the true MTD, which is defined by the percentage of simulated trials that select a dose lower than the true MTD at the end of the trial across all the simulated trials.
- Patient Allocation
  - Percentage of Correct Allocation (PCA): The average percentage of patients who are correctly assigned to the true MTD by the design across all the simulated trials.
  - Percentage of Overdosing Allocation (POA): The average percentage of patients who are assigned to doses higher than the MTD by the design across all the simulated trials.
  - Percentage of Underdosing Allocation (PUA): The average percentage of patients who are assigned to doses lower than the MTD by the design across all the simulated trials.

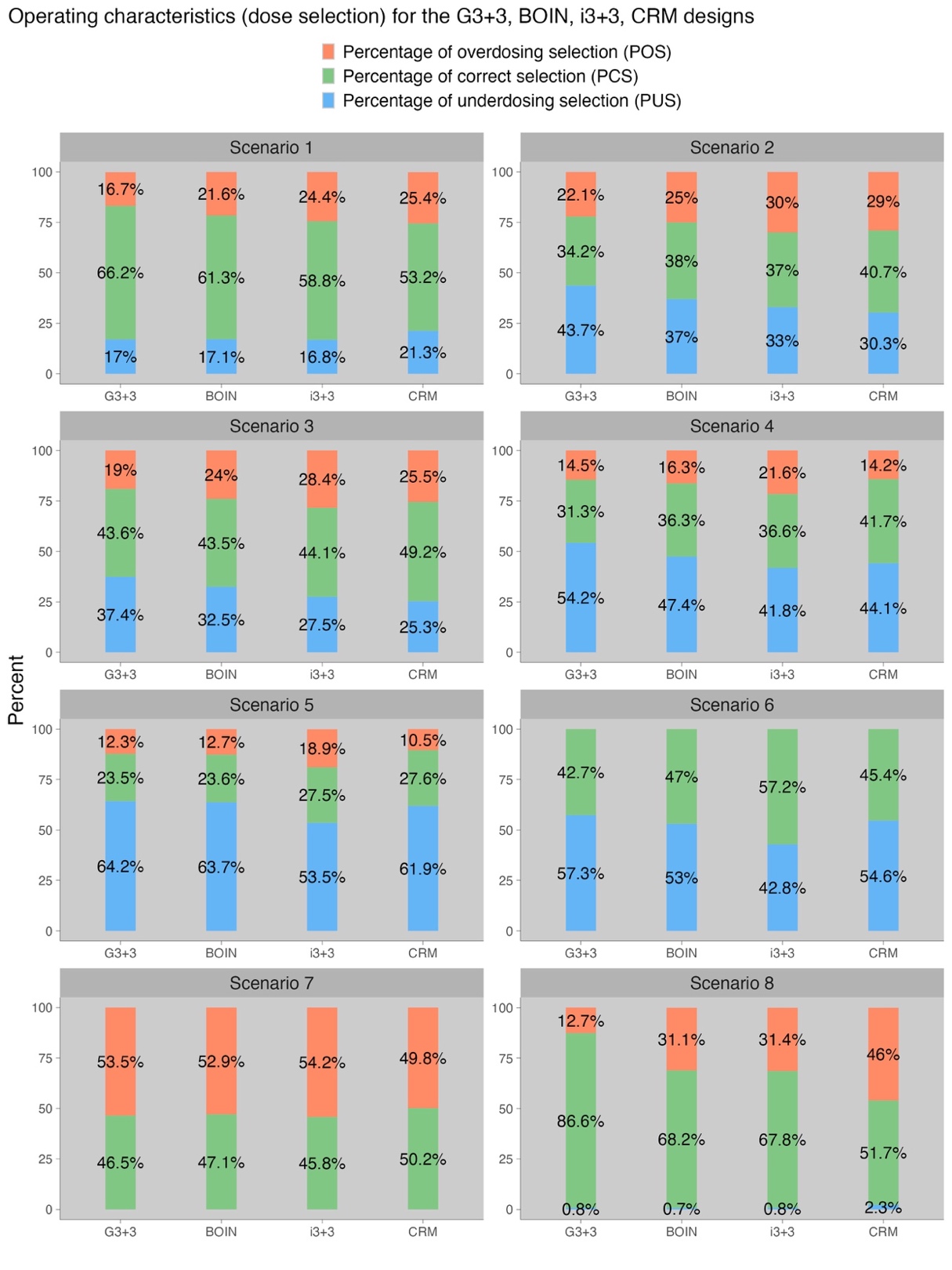
Figure A.2. Operating characteristics in terms of MTD selection for the G3+3, BOIN, i3+3, and CRM designs.

Figure A.3. presents patient allocation results. The G3+3 design is comparable to the BOIN and i3+3 designs. Even though the POA value of G3+3 is slightly larger than the BOIN design, it is due to the fact that the G3+3’s decision is S when $n=3$ and $y=1$ compared with D for BOIN.

Figure A.3. Operating characteristics in terms of patient allocation for the G3+3, BOIN, i3+3 and CRM designs.

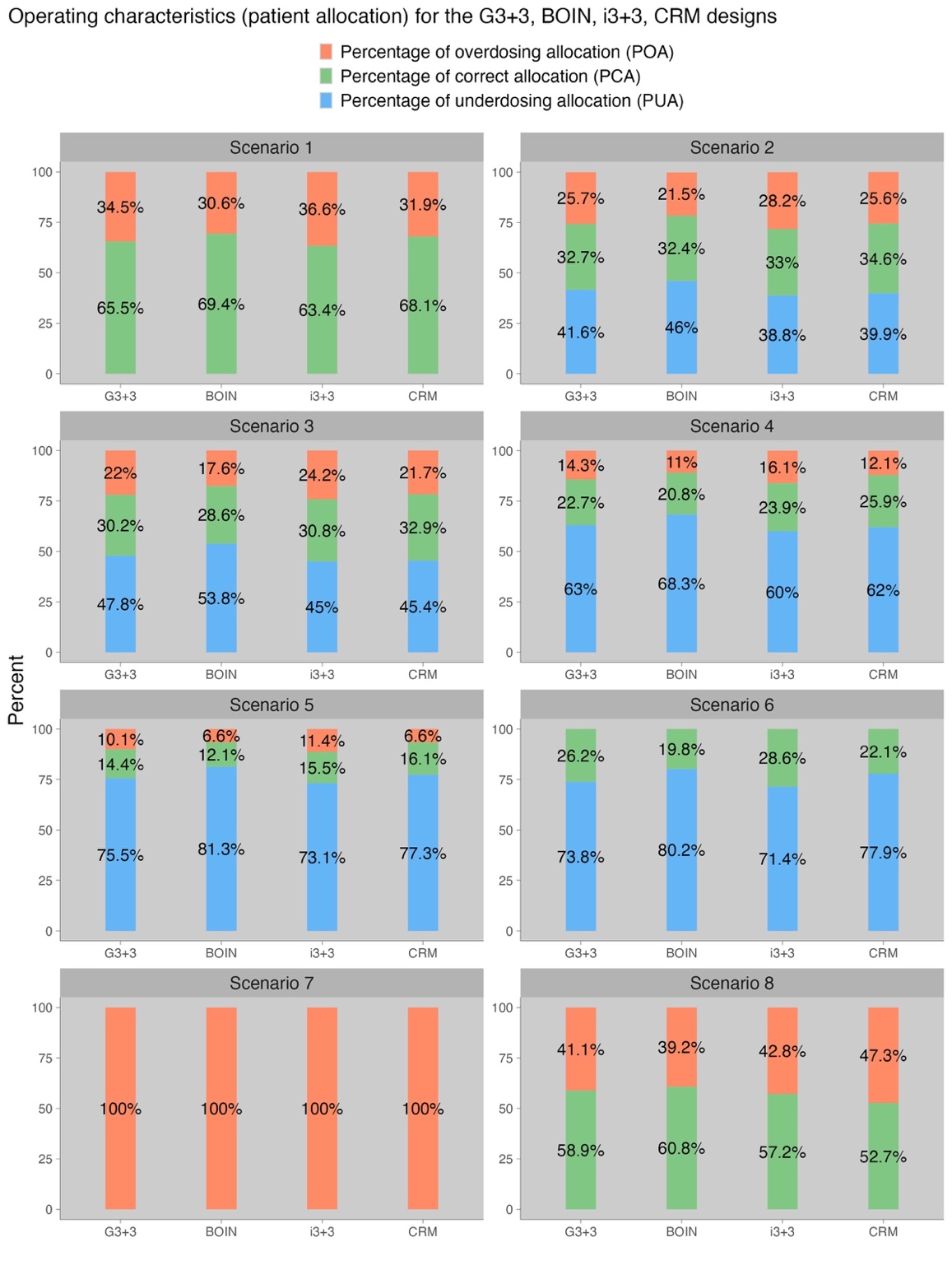

1. These numbers may change for a specific trial.
